# Correlation of post-vaccination fever with specific antibody response to SARS-CoV-2 BNT162b2 booster and no significant influence of antipyretic medication

**DOI:** 10.1101/2022.07.25.22277569

**Authors:** Naoki Tani, Hideyuki Ikematsu, Takeyuki Goto, Kei Gondo, Takeru Inoue, Yuki Yanagihara, Yasuo Kurata, Ryo Oishi, Junya Minami, Kyoko Onozawa, Sukehisa Nagano, Hiroyuki Kuwano, Koichi Akashi, Nobuyuki Shimono, Yong Chong

**Affiliations:** Medicine and Biosystemic Science, Kyushu University Graduate School of Medical Sciences, Fukuoka, Japan; Ricerca Clinica Co., Fukuoka, Japan; Clinical Laboratory, Fukuoka City Hospital, Fukuoka, Japan; Pharmacy, Fukuoka City Hospital, Fukuoka, Japan; Department of Infectious Diseases, Fukuoka City Hospital, Fukuoka, Japan; Department of Neurology, Fukuoka City Hospital, Fukuoka, Japan; Fukuoka City Hospital, Fukuoka, Japan; Center for the Study of Global Infection, Kyushu University Hospital, Fukuoka, Japan

**Keywords:** Antipyretic, SARS-CoV-2, Vaccine, Antibody, Reactogenicity

## Abstract

**Background:** A SARS-CoV-2 mRNA vaccine booster elicits sufficient antibody responses that protect against COVID-19, whereas adverse reactions such as fever have been commonly reported. Associations between adverse reactions and antibody responses have not been fully characterized, nor has the influence of antipyretic use.

**Methods:** This is a prospective observational cohort study in Japan, following our prior investigation of BNT162b2 two-dose primary series. Spike-specific IgG titers were measured for SARS-CoV-2-naive hospital healthcare workers who received a BNT162b2 booster. The severity of solicited adverse reactions, including the highest body temperature, and self-medicated antipyretics were reported daily for seven days following vaccination through a web-based self-reporting diary.

**Results:** The data of 281 healthcare workers were available. Multivariate analysis extracted fever after the booster dose (beta=0.305, p<0.001) as being significantly correlated with the specific IgG titers. The analysis of 164 participants with data from the primary series showed that fever after the second dose was associated with the emergence of fever after the booster dose (relative risk: 3.97 [95% confidence interval: 2.48-6.35]); however, the IgG titers after the booster dose were not affected by fever after the second dose. There were no significant differences in the IgG titers by the use, type, or dosage of antipyretic medication.

**Conclusions:** These results suggest an independent correlation between mRNA vaccine-induced specific IgG levels and post-booster vaccination fever, without any significant influence of fever after the primary series. Antipyretic medications for adverse reactions would not interfere with the elevation of specific IgG titers.

**summary:** Spike-specific IgG titers after a BNT162b2 booster were measured for healthcare workers. Adverse reactions and self-medicated antipyretics were reported. Post-booster vaccination fever was correlated with the specific IgG titers. Antipyretics used for adverse reactions did not suppress specific IgG induction.

## 1. Introduction

Administration of an mRNA coronavirus disease 2019 (COVID-19) vaccine has shown high vaccine efficacy, substantially reducing the risk of severe acute respiratory syndrome coronavirus 2 (SARS-CoV-2) infection and the development of severe or critical disease [1–4]. These efficacy data are consistent with evidence from immunogenicity studies that show robust specific antibody responses to the mRNA COVID-19 vaccines [5,6]. It is notable that the specific IgG and neutralization titers of vaccinees who received an mRNA COVID-19 vaccine exceeded those of recovered COVID-19 patients [7,8]. On the other hand, the reactogenicity of mRNA COVID-19 vaccines is known to be relatively high, and fever is a common adverse reaction. Around 15% of vaccinees had fever after the second or third dose of an mRNA COVID-19 vaccine [1,9,10], while only 1-2% did so after influenza or pneumococcal vaccination [11,12]. The possible relation between adverse reactions, including fever, and the antibody responses to mRNA COVID-19 vaccines remains to be fully elucidated. Antipyretic or pain medications (antipyretics) are often used to mitigate the frequent adverse events. Public health authorities allow the therapeutic use of antipyretics [13,14], but data for the possible influence of their use on the antibody responses to COVID-19 vaccines are insufficient.

We previously investigated the influence of adverse reactions and the use of antipyretics on the specific antibody responses to the two-dose primary series of BNT162b2 vaccine (Pfizer, Inc., and BioNTech). A positive correlation of degree of fever after the second dose and little interference from the antipyretic medications on the antibody titers were shown [15]. In the present study of the same cohort, we prospectively investigated the influence of adverse reactions, which were evaluated using a standardized assessment tool, and the use of antipyretics on the specific antibody responses to a booster dose of BNT162b2 vaccine. In addition, using the data of the participants for whom information on both the second and booster doses were available, we also evaluated whether the specific antibody titers after the booster dose are affected by fever after the second dose.

## 2. Participants and methods

### 2.1. Participants

Eligible participants were healthcare workers who received three 30µg doses of BNT162b2 at Fukuoka City Hospital in Japan. The primary two-dose vaccine series with a 21-day interval was administered between March and June 2021, and the booster dose was given between December 2021 and January 2022. Included in the analysis were vaccinees who had serum sampling done 14 days or more after the booster dose and who completely responded to questionnaires about their background and solicited adverse reactions. The exclusion criteria were: 1) previously diagnosed with COVID-19 by laboratory tests, 2) positive results for antibodies targeting the viral nucleocapsid protein (IgG(N)), 3) the use of an antipyretic within 24 hours before the booster dose, and 4) receiving immunosuppressive therapy.

All participants provided written informed consent before undergoing any of the study procedures. The study was approved by the ethics review board of Fukuoka City Hospital (approval number 228) and registered in the University Hospital Medical Information Network-Clinical Trials Registry (registration no. UMIN000046246).

### 2.2. Demographic characteristics, reactogenicity, and antipyretic medications

Participant background information was collected by a web-based questionnaire. Local and systemic adverse reactions were reported daily for seven days after the booster dose through a web-based self-reporting diary. The solicited data were as follows: 1) local reactions (pain at the infection site, redness, and swelling), and 2) systemic events (fever, fatigue, headache, chills, vomiting, diarrhea, muscle pain, joint pain, and lymphadenopathy). Axillary body temperature was measured twice daily, morning and night, and whenever the participant felt feverish. The highest body temperature during the seven days was used in the analysis. All solicited reactions except lymphadenopathy were recorded based on standardized assessment scales developed by the US Food and Drug Administration guidelines [16].

Lymphadenopathy was evaluated for its presence or absence. The use of an antipyretic was left up to the participant. Information on the self-medicated antipyretics, including name, dosage, timing, and reason for use, was collected daily with the solicited adverse reactions for the seven days after the booster dose.

Previously collected data on the IgG(S-RBD) titers and adverse reactions to the second dose were used in the present study. The major differences in the method of data collection were that in the earlier studies the adverse reaction information was collected for five days, not seven, after each of the doses and that we had used an originally defined subjective scaling method, except for fever. The detailed methods are shown in our previous study [15].

### 2.3. Serological testing

Serum samples were collected twice, before and after the booster dose. The interval between the booster dose and the sampling after vaccination was scheduled at approximately one month to match it with the interval between the second dose and the sampling [15]. The quantitative levels of IgG antibodies for the receptor binding domain of the S1 subunit of the viral spike protein (IgG(S-RBD)) and IgG(N) were measured using the SARS-CoV-2 IgG II assay and SARS-CoV-2 IgG assay, respectively (Abbott Laboratories Co., Ltd., Park, IL, USA). Signal-to-cutoff values of ≥1.4 AU/mL were applied for IgG(N) positivity [17].

### 2.4. Statistical analysis

The IgG(S-RBD) titers were log-transformed for analysis. The median, interquartile range (IQR), geometric mean titer (GMT), fold change, 95% confidence interval (CI), and relative risk (RR) were calculated. Between-group differences were calculated with Student’s t test, ANOVA, or post hoc Dunnett’s test in line with suitability. Correlation coefficients were calculated using Spearman’s correlation coefficient. Multivariate linear regression models with a stepwise selection procedure were established. The level of significance was set at <5%, two-sided. All analyses were performed using the SAS software package, release 9.4 (SAS Institute, Cary, NC).

## 3. Results

### 3.1. Demographic characteristics

Among 419 staff members who received the BNT162b2 booster, serum samples were collected from 346, and 316 satisfied the inclusion criteria. Of these, 13 were excluded due to a history of COVID-19 or IgG(N) ≥1.4 AU/mL, 20 due to pre-vaccination use of antipyretics, and 2 due to having received immunosuppressive therapy, leaving the data of 281 participants available for analysis. Demographic and background information are summarized in Table 1. The median age was 41 years (IQR, 33-50), 72.6% were female, all were Japanese, and 77.2% had no coexisting conditions. The median interval between the second and booster doses was 262 days (IQR, 260-264, range, 219-288). Serum samplings before and after the booster dose were taken approximately eight months after the second dose (median, 247 days, IQR, 244-252) and one month after the booster dose (median, 32 days, IQR, 29-33), respectively.

**Table 1:**
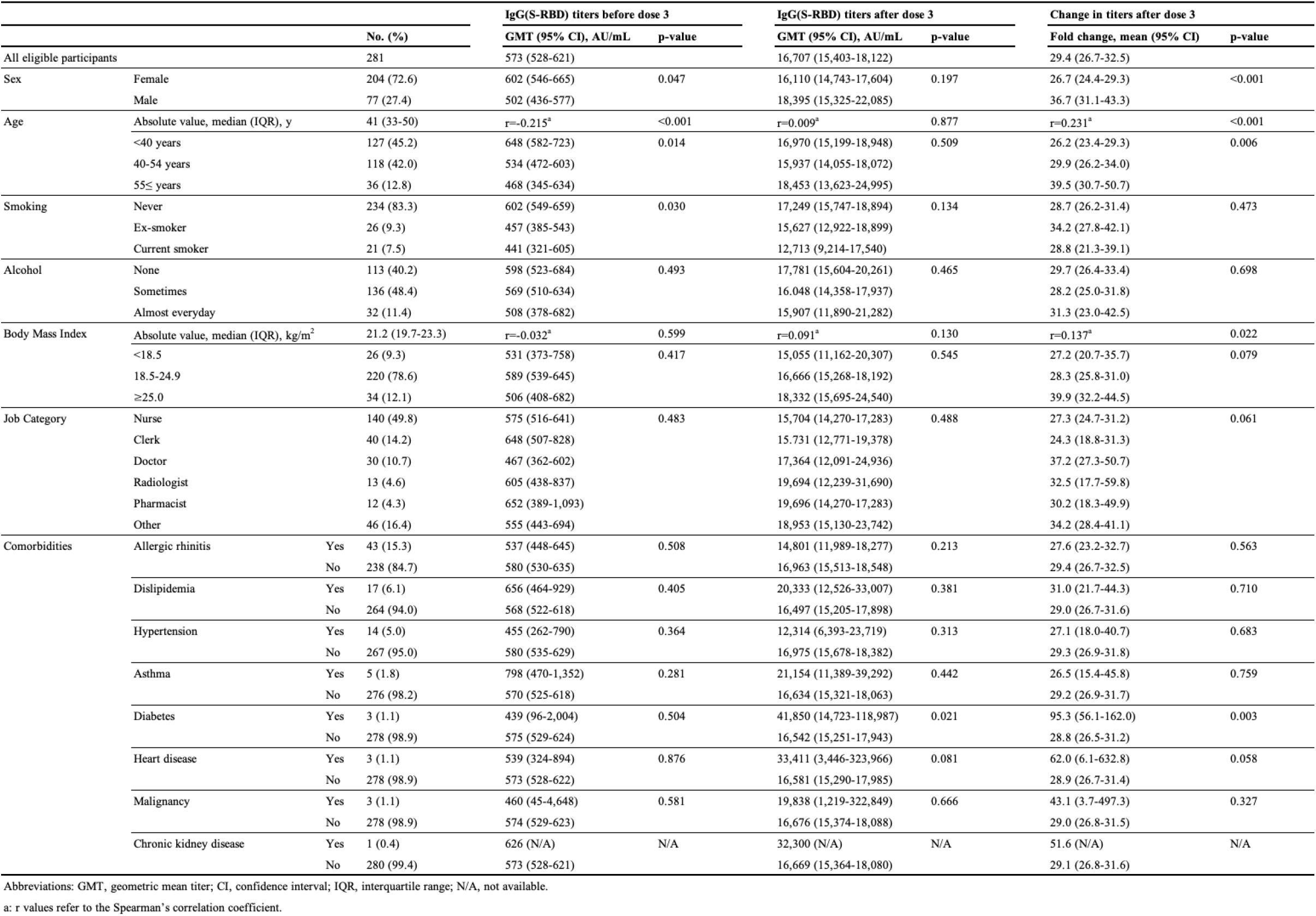
Demographic characteristics, IgG(S-RBD) titer, and fold change.

### 3.2. IgG(S-RBD) titer and fold change by demographic characteristics

The booster dose increased the GMT of IgG(S-RBD) 29.4-fold (95% CI, 26.7-32.5), from 573 AU/mL (95% CI, 528-621) to 16,707 AU/mL (95% CI, 15,403-18,122). The IgG(S-RBD) titers and fold changes according to demographic characteristics are shown in Table 1. In the analysis of the IgG(S-RBD) titers, sex and age showed no significant correlation (p=0.197 and p=0.645, respectively). In contrast, both were significantly correlated with the fold changes. The mean fold-change in males was higher than that in females (36.7-fold (95% CI, 31.1-43.3) vs 26.7-fold (95% CI, 24.4-29.3), p<0.001). Age showed a positive correlation with the fold changes (r=0.244, p<0.001). Comorbidities other than diabetes were not correlated. Diabetes had statistically significant correlations with both the IgG(S-RBD) titers and fold changes, but it was not further analyzed due to the small number of participants with diabetes.

### 3.3. IgG(S-RBD) titer and fold change by adverse reaction

The IgG(S-RBD) titers and fold changes after the booster dose by the solicited adverse reactions are shown in Table 2. None of the local reactions had a significant influence on the IgG(S-RBD) titers. Among the systemic reactions, fever showed a positive correlation with the IgG(S-RBD) titers (r=0.262, p<0.001). Fatigue (p=0.022), headache (p=0.024), chills (p=0.001), and lymphadenopathy (p<0.001) were also positively correlated. Neither local nor systemic adverse reactions significantly affected the fold changes in IgG(S-RBD) titers.

**Table 2:** Influence of adverse reaction variables on JgG(S-RBD) titer.

|  |  | No. (%) | GMT (95% CI), AU/mL | p-value | Fold change, mean (95% CI) | p-value |
| --- | --- | --- | --- | --- | --- | --- |
| **Local reactions** |  |  |  |  |  |  |
| Pain at injection site | No | 3 (1.1) | 14,818 (4,032-54,450) | 0.763 | 23.9 (10.9-52.4) | 0.618 |
|  | Yes | 278 (98.9) | 16,730 (15,413-18,155) |  | 29.2 (26.9-31.7) |  |
| Redness | No | 185 (65.8) | 16,761 (15,198-18,484) | 0.915 | 28.9 (26.2-31.9) | 0.734 |
|  | Yes | 96 (34.2) | 16,604 (14,328-19,244) |  | 29.7 (25.7-34.4) |  |
| Swelling | No | 148 (52.7) | 16,707 (14,866-18,772) | 0.998 | 30.5 (27.4-33.8) | 0.269 |
|  | Yes | 133 (47.3) | 16,707 (14,907-18,728) |  | 27.8 (24.5-31.6) |  |
| **Systemic reactions** |  |  |  |  |  |  |
| Fever | Absolute value | 281 (100) | r=0.262<sup>a</sup> | <0.001 | r=0.082<sup>a</sup> | 0.170 |
|  | <37.0°C | 114 (40.6) | 14,011 (12,431-15,793) | <0.001 | 26.9 (23.7-30.5) | 0.100 |
|  | 37.0-37.9°C | 97 (34.5) | 16,302 (14,174-18,750) |  | 28.8 (25.3-32.9) |  |
|  | ≥38.0°C | 64 (24.9) | 23,020 (19,670-26,941) |  | 33.7 (28.1-40.5) |  |
| Fatigue | No | 42 (15.0) | 13,329 (10,713-16,581) | 0.022 | 26.2 (20.8-32.9) | 0.278 |
|  | Yes | 239 (85.1) | 17,382 (15,933-18,967) |  | 29.7 (27.2-32.4) |  |
| Headache | No | 94 (33.5) | 14,652 (12,885-16,661) | 0.024 | 29.3 (25.6-33.5) | 0.947 |
|  | Yes | 187 (66.6) | 17,848 (16,099-19,783) |  | 29.1 (26.3-32.3) |  |
| Chills | No | 135 (48.0) | 14,534 (13,077-16,151) | 0.001 | 27.3 (24.3-30.7) | 0.126 |
|  | Yes | 146 (52.0) | 19,006 (16,862-21,419) |  | 31.0 (27.6-34.8) |  |
| Vomiting | No | 272 (96.8) | 16,512 (15,198-17,943) | 0.121 | 29.1 (26.7-31.6) | 0.721 |
|  | Yes | 9 (3.2) | 23,752 (16,021-35,221) |  | 31.6 (22.5-44.5) |  |
| Diarrhea | No | 246 (87.5) | 16,811 (15,385-18,365) | 0.693 | 29.3 (26.8-32.0) | 0.812 |
|  | Yes | 35 (12.5) | 15,999 (13,005-19,683) |  | 28.4 (23.6-34.2) |  |
| Muscle pain | No | 40 (14.2) | 17,434 (14,142-21,493) | 0.674 | 28.9 (22.2-37.7) | 0.944 |
|  | Yes | 241 (85.8) | 16,588 (15,181-18,126) |  | 29.2 (26.8-31.8) |  |
| Joint pain | No | 128 (45.6) | 16,044 (14,315-17,980) | 0.370 | 27.9 (24.6-31.6) | 0.335 |
|  | Yes | 153 (54.5) | 17,282 (15,396-19,404) |  | 30.2 (27.1-33.7) |  |
| Lymphadenopathy | No | 194 (69.0) | 15,332 (13,845-16,975) | <0.001 | 29.1 (26.1-32.4) | 0.956 |
|  | Yes | 87 (31.0) | 20,234 (17,857-22,935) |  | 29.2 (26.2-32.7) |  |

Abbreviations: GMT, geometric mean titer; CI, confidence interval; IQR, interquartile range; N/A, not available.
a: r values refer to the Spearman's correlation coefficient.

Factors that showed a p-value of <0.2, including sex, body mass index, smoking history, heart disease, fever, fatigue, headache, chills, vomiting, and lymphadenopathy, in the univariate analyses were incorporated in multivariate analysis. Among these variables, only fever (p<0.001, standardized regression coefficient beta 0.305 [95% CI, 0.193-0.417]) was extracted as being independently correlated with the IgG(S-RBD) titers (adjusted R^2^ 0.090). A similar analysis for the fold change in IgG(S-RBD) titers extracted age (p<0.001, beta 0.248 [95% CI, 0.136-0.360]), male sex (p=0.001, beta 0.183 [95% CI, 0.071-0.294]), and fever (p=0.018, beta 0.136 [95% CI, 0.024-0.248]) as significant (adjusted R^2^ 0.105).

### 3.4. Influence of antipyretic medications on IgG(S-RBD) titer and fold change

The analyses of the influence of antipyretics on the IgG(S-RBD) titers and fold changes are shown in Table 3. In total, 119 (42.4%) participants used antipyretics during the seven days after the booster dose. None of the participants prophylactically used an antipyretic after the vaccination to prevent adverse events. The GMT of IgG(S-RBD) was comparable between the groups with and without antipyretic use (17,466 AU/mL (95% CI, 15,279-19,966) vs 16,170 AU/mL (95% CI, 14,601-17,906), p=0.357). Similarly, there was no significant difference in the mean fold-change between the two groups (29.4-fold (95% CI, 26.1-33.2) vs 28.9-fold (95% CI, 25.9-32.4), p=0.893). No significant influence of antipyretic use was found when stratified by fever grade, classified into <37.0 °C, 37.0-37.9 °C, and ≥38.0 °C. The most commonly used antipyretic combination was acetaminophen monotherapy (43/119, 36.1%), followed by loxoprofen monotherapy (35/119, 29.4%) and ibuprofen monotherapy (14/119, 11.8%). The IgG(S-RBD) titers and fold changes of the group with non-steroidal anti-inflammatory drugs (NSAIDs) such as loxoprofen and ibuprofen were comparable to those of acetaminophen. The IgG(S-RBD) titers by all combinations of antipyretics used are summarized in Table s1.

**Table 3:**
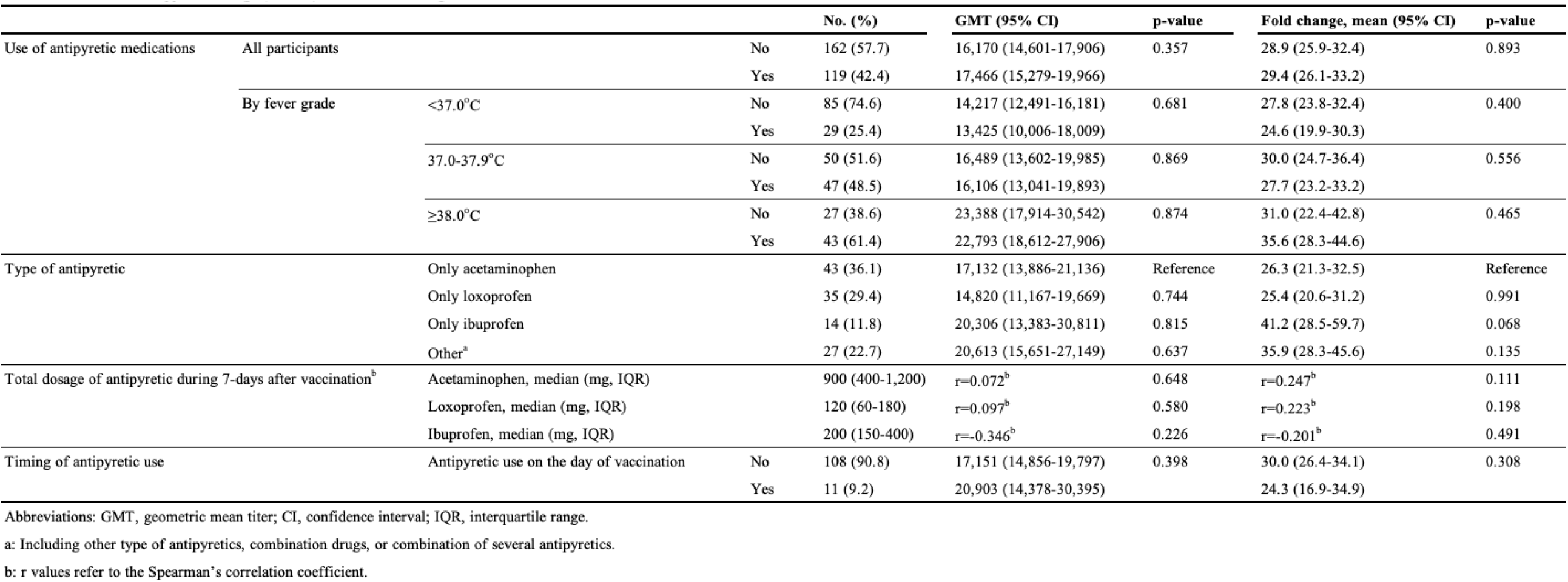
Influence of antipyretics on lgG(S-RBD) titer and fold change after a booster dose.

### 3.5. Association between the presence or absence of an adverse reaction after the second dose and the emergence of the same reaction after the booster dose

Using the data of 164 participants for whom information on both the second and booster doses were available, the probability of a solicited adverse reaction after the booster dose was investigated by the presence or absence of the corresponding reaction after the second dose (Table 4). The RR of all solicited adverse reactions were over 1.0. The presence of swelling at the infection site, fever of 38.0 ≥oC, headache, chills, muscle pain, and joint pain after the second dose showed a significant association with the emergence of the corresponding reaction after the booster dose. The RR was the highest for the fever, at 3.97 (95% CI, 2.48-6.35).

**Table 4:** Probability of an adverse reaction after the booster dose according to the presence or absence of the same reaction after the second dose.

|  |  | Corresponding reactions after booster dose |  | Incidence of a corresponding reaction<br>after booster dose, % | RR | 95% CI |
| --- | --- | --- | --- | --- | --- | --- |
|  |  | Present | Absent |  |  |  |
| Local reactions after dose 2 |  |  |  |  |  |  |
| Pain at injection site | Present | 154 | 1 | 99.4 | 1.12 | 0.89-1.41 |
|  | Absent | 8 | 1 | 88.9 | Reference | Reference |
| Redness | Present | 14 | 16 | 46.7 | 1.56 | 0.98-2.48 |
|  | Absent | 40 | 94 | 29.9 | Reference | Reference |
| Swelling | Present | 45 | 23 | 66.2 | 1.93 | 1.39-2.66 |
|  | Absent | 33 | 63 | 34.4 | Reference | Reference |
| Systemic reactions after dose 2 |  |  |  |  |  |  |
| Fever $\geq 38.0^{\circ}\text{C}$ | Present | 21 | 12 | 63.6 | 3.97 | 2.48-6.35 |
|  | Absent | 21 | 110 | 16.0 | Reference | Reference |
| Fatigue | Present | 129 | 14 | 90.2 | 1.35 | 1.00-1.84 |
|  | Absent | 14 | 7 | 66.7 | Reference | Reference |
| Headache | Present | 80 | 19 | 80.8 | 1.46 | 1.15-1.85 |
|  | Absent | 36 | 29 | 55.4 | Reference | Reference |
| Chills | Present | 52 | 18 | 74.3 | 2.00 | 1.48-2.68 |
|  | Absent | 35 | 59 | 37.2 | Reference | Reference |
| Diarrhea | Present | 3 | 17 | 15.0 | 1.14 | 0.37-3.50 |
|  | Absent | 19 | 125 | 13.2 | Reference | Reference |
| Muscle Pain | Present | 85 | 6 | 93.4 | 1.22 | 1.06-1.40 |
|  | Absent | 56 | 17 | 76.7 | Reference | Reference |
| Joint Pain | Present | 59 | 21 | 73.8 | 1.77 | 1.33-2.35 |
|  | Absent | 35 | 49 | 41.7 | Reference | Reference |

Abbreviations: RR, relative risk; CI, confidence interval.
Information on vomiting and lymphadenopathy after the second dose were not collected.

### 3.6. IgG(S-RBD) titer by the presence of fever after the second and booster doses

The 164 participants were divided into four groups by the presence or absence of fever of ≥38.0°C after each dose. The IgG(S-RBD) titers of these four groups one month after the second dose, eight months after the second dose, and one month after the booster dose are shown in Table 5. The GMTs of IgG(S-RBD) one month after the second dose were higher for the groups with fever after the second dose than for those without. Eight months after the second dose, the differences among the groups were subtle, ranging from 537 to 796 AU/mL, without significant differences. The GMTs of IgG(S-RBD) one month after the booster dose were higher in the groups with fever after the booster dose, and they were comparable between the groups with and without fever after the second dose. A multivariate analysis incorporating fever after the second dose was additionally done. Fever after the booster dose was extracted as being significantly correlated with the IgG(S-RBD) titers after the booster dose (p=0.001, beta 0.246 [95% CI, 0.097-0.395]), but fever after the second dose was not.

**Table 5.** IgG(S-RBD) titers by the presence or absence of fever after the second and booster doses.

| Fever $\geq 38.0^{\circ}\text{C}$ after vaccination<br>(after dose 2/dose 3) | No. (%) | Female sex (%) | Age, median (IQR), y | 1-month after dose 2 | | 8-months after dose 2 | | 1-month after dose 3 | |
| --- | --- | --- | --- | --- | --- | --- | --- | --- | --- |
|  |  |  |  | GMT (95% CI), AU/mL | p-value | GMT (95% CI), AU/mL | p-value | GMT (95% CI), AU/mL | p-value |
| Absent/Absent | 110 (67.1) | 83 (75.5) | 42.0 (33.0-49.0) | 8,281 (7,305-9,386) | Reference | 537 (474-607) | Reference | 15,438 (13,665-17,441) | Reference |
| Present/Absent | 12 (7.3) | 9 (75.0) | 33.5 (29.0-39.5) | 12,026 (8,204-17,628) | 0.169 | 796 (552-1,147) | 0.144 | 14,644 (10,630-20,174) | 0.990 |
| Absent/Present | 21 (12.8) | 12 (57.1) | 41.0 (46.0-51.0) | 9,172 (6,717-12,524) | 0.879 | 738 (515-1,058) | 0.125 | 22,012 (16,555-29,270) | 0.060 |
| Present/Present | 21 (12.8) | 17 (81.0) | 40.0 (28.0-44.0) | 12,343 (9,562-15,932) | 0.032 | 627 (481-817) | 0.682 | 22,479 (16,575-30,487) | 0.042 |

Abbreviations: IQR, interquartile range; GMT, geometric mean titer; CI, confidence interval.

## 4. Discussion

A booster dose of an mRNA COVID-19 vaccine induces a higher antibody titer than is produced by the primary two-dose series [18,19]. Our previous study showed that the IgG(S-RBD) titers after the second dose of the primary series were relatively low for male participants and the elderly [15], as shown in other studies [20,21]. In contrast, in the present study of the booster dose for the same cohort, male sex and age showed positive correlations with the fold change in IgG(S-RBD) titers, reaching comparable levels in the IgG(S-RBD) titers by sex and age. Other studies have also shown that the specific IgG titers after the BNT162b2 booster were comparable for sex and age [22,23]. The mechanism for the difference in the immunogenicity of a BNT162b2 vaccine by sex and age between the second and booster doses is unclear, but the booster vaccination would drive antibody productions, especially in the populations with relatively weak responses to the primary series.

The relation between the emergence of vaccine-related adverse reactions and antibody induction by the primary series of mRNA COVID-19 vaccines has been reported. Studies that used scores based on the sum of the presence or a severity scale of solicited adverse reactions showed no significant correlation of adverse reactions with the spike-specific IgG titers [24–26]. On the other hand, when each reaction was separately analyzed, the presence of fever after the second dose was shown to be correlated with high IgG titers [27]. In the study, fever was used in the analysis by its presence or absence, not degree. We previously showed that degree of fever after the second dose was correlated with the IgG(S-RBD) titers by a multivariate analysis [15]. In the present study, the degree of fever after the booster dose, which was defined by the highest body temperature after the vaccination, was again shown to have a significant correlation with the IgG(S-RBD) titers. The mechanism of how fever after SARS-CoV-2 mRNA vaccination is linked to the antibody production remains unclear. Elucidating the mechanism will lead to a better understand of the sufficient antibody induction mechanism of mRNA COVID-19 vaccines.

The possibility of a relation between post-vaccination fever and the SARS-CoV-2 specific antibody titers has created concern that antipyretics, which can suppress fever, may have a negative influence on antibody responses to SARS-CoV-2 vaccination. To date, few studies have been done to determine the influence of antipyretics on the immunogenicity of COVID-19 vaccines. We previously showed that therapeutic use of antipyretics did not interfere with the antibody responses to the primary BNT162b2 series [15]. In the present study for the BNT162b2 booster, no influence of antipyretic use on the IgG(S-RBD) titers was observed again. Although several in vitro laboratory studies have demonstrated that NSAIDs inhibit several pathways leading to antibody responses [28–30], the IgG(S-RBD) titers of the group with NSAIDs were comparable to those of the group with acetaminophen. The present study was not designed to evaluate the influence by the type of antipyretics, but it is suggested that neither acetaminophen nor NSAIDs would interfere with the elevation of IgG(S-RBD) titers. Antipyretics may also be used prophylactically. One study of a COVID-19 adenoviral vector vaccine reported that prophylactic acetaminophen use reduced many adverse reactions without interfering with antibody responses [31]. There are no studies on the influence of prophylactic use of antipyretics on antibody responses to mRNA COVID-19 vaccines. Taken together, our results indicate that the therapeutic use of antipyretics, regardless of the type, acetaminophen or NSAIDs, would be helpful for alleviating adverse reactions, including fever, without interfering with antibody responses to SARS-CoV-2 mRNA vaccination.

It would be of great interest for physicians whether the presence of an adverse reaction after the two-dose primary series is useful to predict the emergence of the corresponding reaction after the booster dose. In the present study, the presence of swelling as a local reaction and several systemic reactions including fever after the second dose was associated with the emergence of the corresponding reactions after the booster dose. Fever after the second dose showed the highest RR of 3.97. Of interest, fever after the booster dose was independently correlated with the IgG(S-RBD) titers after the booster dose, irrespective of fever after the second dose. Thus, the participants with fever after the second dose may be more likely to have fever after the booster dose, but fever after the second dose would not affect the antibody responses to the booster dose. To our knowledge, no investigations of the correlation between post-vaccination fever and specific IgG levels have been done in the same cohort throughout the primary and booster vaccinations. These findings impress a potential linking of post-vaccination fever with the antibody production induced by mRNA COVID-19 vaccines.

This study has some limitations that should be acknowledged. First, it is a single-center observational study with a relatively small sample size of 281 participants. Additionally, the population was relatively young and predominantly female. Second, the data collection methods for the solicited adverse reactions were slightly different between the second and booster doses. The data collection period for adverse reactions was seven days after vaccination for the booster dose, while it was five days for the second dose. However, most common adverse reactions have been known to occur within a few days after vaccination [32], thus, the difference would have little impact on our findings. Finally, the type, dosage, and timing of antipyretic usage were chosen by each participant and are thus arbitrary. Randomized, large scale, controlled studies will be necessary to clarify the influence of antipyretics on the immunogenicity outcomes of COVID-19 vaccines, but we feel that our real-world data from healthcare workers with self-medicated antipyretics is informative.

In conclusion, a booster dose of BNT162b2 vaccine can restore waning immunity and significantly increase the antibody titers, especially in males and the elderly who had relatively low antibody responses to the primary two-dose series. Post-booster vaccination fever could be independently correlated with mRNA vaccine-induced specific IgG levels, without any significant influence of fever after the primary series. Antipyretic medications would be helpful to alleviate suffering from adverse reactions, without suppressing the specific IgG responses induced by a BNT162b2 vaccine booster.

## Data Availability

All data produced in the present study are available upon reasonable request to the authors

## Acknowledgments

We thank all of the staff members of Fukuoka City Hospital for their support for sample collection and data reduction.

